# Understanding frailty: probabilistic causality between components and their relationship with death through a Bayesian network and evidence propagation

**DOI:** 10.1101/2022.03.02.22271711

**Authors:** Ricardo Ramírez-Aldana, Lorena Parra-Rodríguez, Juan Carlos Gomez-Verjan, Carmen Garcia-Peña, Luis Miguel Gutierrez-Robledo

## Abstract

Identifying relationships between components of an index helps to have a better understanding of the condition they define. The Frailty Index (FI) measures the global health of individuals and can be used to predict outcomes as mortality. Previously, we modelled the relationship between the FI components (deficits) and death through an undirected graphical model, analyzing their relevance from a social network analysis framework. Here, we model the FI components and death through an averaged Bayesian network obtained through a structural learning process and resampling, to understand how the FI components and death are causally related. We identified that the components are not similarly related between them and that deficits are related according to its type. Two deficits were the most relevant in terms of their connections and other two were directly associated with death. We also obtained the strength of the relationships to identify the most plausible, identifying clusters of deficits heavily related. Finally, we propagated evidence (assigned values to all deficits) and studied how FI components predict mortality, obtaining a correct assignation of almost 74%, whereas a sensitivity of 56%. As a classifier of death, the more number of deficits included for the evidence, the best performance; but the FI seems not to be very good to correctly classify death people.

## Introduction

Frailty is a term widely used to denote a state of poor homeostatic resolution (vulnerability) after a stressor event and is a consequence of loss of homeostatic reserve across multiple physiological systems, which occurs across the lifetime [8]. Frailty appears to be a valid construct, since it can be used to identify older adults who are at increased risk of mortality and functional decline when they are hospitalized [3], but consensus has yet to be reached on how exactly to define and determine it, so we could make this term useful for clinics.

One approach to measure frailty is to count deficits in health (symptoms, signs, diseases, disabilities or laboratory, radiographic, or electrocardiographic abnormalities) on the grounds that the more deficits a person has, the more likely that such individual could be frail [44,50].

Variables included in the frailty index (FI) satisfy that they are associated with health status and that their prevalence must generally increase with age, but do not become too prevalent at some younger age. The deficits that make up the FI cover a range of systems; and if a single FI is used serially on the same people, the items that make up the FI need to be the same from one iteration to the next [50]. A robust FI requires a significant number of individual items (between 30 and 40), utilized to record deficit accumulation, and which are recorded as a score or index.

The FI approach does not assume that the deficits that make up frailty are statistically independent [46], and usually includes comorbidity (the concurrent presence of two or more medically diagnosed diseases) and disability (difficulty or dependency in performing activities of daily living) variables [45]. It is still unclear if disabilities and comorbidities themselves should be a domain to define frailty or if they already constitute adverse health outcomes [30, 57] and how health deficits connect and interact with each other and to how the FI is associated with death, although several theoretical complex network models have been proposed [13-14, 17, 36, 47, 56].

On the other hand, a Bayesian network is a complex model that allows to identify independences and causal relationships between variables, considering all variables at a same level, in the sense that there is not necessarily a division of them as explanatory and response variables. Additionally, we can see how a subset of variables are affected conditional to values provided to other subset of variables also known as evidence propagation. Thus, this methodology simultaneously allows to perform explanatory and predictive analyses. Therefore, in the present work, we analyzed frailty and its components through this technique, which allowed us to have a better understanding of how they are working together and with death. Frailty has been analyzed before through simulation methods, for instance through social network analysis models [13, 36, 47, 56]; thus, analyzing frailty using real data seems to be a priority to validate such models. In this context, we have already presented probabilistic graphical models using real data through a Markov network [17], an undirected graphical model; thus, we did not consider directionality, or assigned values to some variables (evidence) and derived through algorithms the probabilities associated with other variables conditional to such assignation (evidence propagation), or the strength of the relationships as we do here.

The main purposes of this work are: 1) To obtain in not simulated data, through a Bayesian network, causal inference (probabilistic causality) concerning the deficits defining the FI and death, seeing whether all variables are similarly related or whether some variables are more related with others, 2) To identify how the variables are related between them and with death considering the strength of the relationships, and 3) To obtain how the probability of death under the model is modified in each individual according to assignation of values (evidence) in certain variables; and, according to this process, to evaluate whether dead and alive people can be correctly classified using an adequate cut-off value for the probability and different sets of variables providing evidence.

## Methods

### Data

The analysis is derived from the longitudinal data from the third and fourth waves (2012 and 2015) from the Mexican Health and Aging Study (MHAS) [62]. MHAS baseline was nationally representative of Mexicans born prior to 1951. A set of questionnaires; socio-demographic, health-related, cognitive performance, functional status, among others, was applied to Mexican older adults over four waves (2001, 2003, 2012, 2015, and 2018). We built all deficits from the 2012 wave and mortality was obtained using information from the 2015 wave and corresponds to deaths occurred between 2012 and 2015. In the 2012 wave, 15,723 participants were assessed, including 9,827 follow-up participants, and 5,896 from a refresh sample (including spouses of the chosen individuals, regardless of age), 1,209 deaths were reported by 2015.

For this study, we used a sample of 14,872 individuals since participants with less than 50 years old were eliminated. Independence between individuals is assumed in the models; thus, correlation analyses were performed finding that correlation between couples was low for all variables, thus couples were included in all analyses. We discarded second or third wives or husbands (165 cases) to avoid problems concerning marital status transitions through time. Though missing data imputation is possible using Bayesian networks, we preferred to discard cases with missing values in any variable to avoid any masking of the sensitivity and specificity associated with the evidence propagation. Hence, from 14,707 cases, a final sample of 10,983 was retained for the present analysis.

### Frailty

A set of 35 variables or deficits, all self-reported, was included as determined by Searle et al. [50]. Variables included concern functional status, chronic diseases, self-rated health, cognitive status, and depressive symptoms. The variables were obtained, categorized, and named as shown in Appendix 1 (supplementary material) and are explained with more detail in [17], where we showed frequencies, calculated the FI for each individual, and showed tables concerning tertiles associated with the index.

### Analytic plan

Bayesian networks (BNs) are discrete probabilistic graphical models [31], which use several concepts defined in graph theory [5]. All variables correspond to vertices in a set V, and the arcs (directed edges) belong to a set A, obtaining a directed graph or digraph D(V,A), which in BNs should be a directed acyclic graph (DAG). The network represents a factorization of the joint probability distribution (probability associated with all variables) corresponding to the random variables associated with each vertex, depending on the graph structure and the Markov properties. These properties state conditional independence relationships between sets of variables. In BNs, the factorization depends on the probability density associated with each variable conditional to the parent nodes (nodes having any directed edge pointing to v). BNs are also used to represent probabilistic causality between sets of variables.

The graph structure that best represents some data or structural learning process is obtained through algorithms (e.g. [22]), some of them are based on optimizing scores, for instance the hill-climbing algorithm. To improve the robustness of the structural learning process, we used a resampling technique, bootstrap method, to learn a set of 100 network structures to use for model averaging [39]. Additionally, to avoid illogical directions between variables, we restricted the structural learning process to avoid arcs from death to any other node (black-list of directions). We obtained the network structure from each bootstrap sample with a hill-climbing search using the Bayesian Information Criterion as score and an empty graph (graph without edges) as initial model. Then, a strength (probability) associated with the presence and direction of each arc is obtained from the bootstrap method.

An average network can be derived by defining a threshold for the strength of both, the presence and direction of the arcs. To determine the presence of arcs, we decided to use a threshold that the algorithm chooses as the one that best represents the data (which in our data was close to 0.5) and to define the direction, we used the most probable direction (strength above or equal to 0.5). Since there are instances in which an arc can have both directions (Stairs to Effort and Effort to Stairs), thus obtaining a network with both directed an undirected edges, we chose the model whose associated DAG pertains to the same equivalence class of the model with the graph having directed and undirected edges. Hence, there could be arcs with a direction strength lower than 0.5 (three in our case, one from “Meds” to “Meals”, another from “Toilet” to “Walk”, and a last one from “Effort” to “Lift”, which originally were in the opposite direction). The DAG associated with the chosen model was drawn using different colors, graphically representing which relationships are stronger.

The parameters (conditional probabilities of a node given its parents) associated with the chosen model were estimated from the data. After that, the network was compiled and since there were not undefined probabilities, evidence propagation was possible [10]. Evidence propagation means that values are provided to a subset of variables, evidence **e**, and its effect over another subset of variables **n** is derived obtaining the conditional probability of the variables in **n** given the evidence, P(**n**|**e**).

We used as evidence the values associated with all variables except death, and propagated it over the death variable, obtaining the probability of death under the model conditional to each combination of values the other variables can take. Then, individuals can be assigned, according to the model and evidence, into dead or alive, by defining a threshold associated with the probability of dying. The default threshold of 0.5 was not appropriate in our data since all probabilities were under 0.5; thus, classifying all dead people as alive. Consequently, we decided to obtain a ROC curve, a plot representing for different thresholds the associated sensitivity, dead people assigned as such in our model, and specificity, alive people assigned as such in our model. We examined the area under ROC curve and chose the threshold maximizing both measures by optimizing the sum of sensitivity and specificity (Youden’s J statistic). To see whether all deficits defining frailty were relevant to determine an accurate probability of death, a similar analysis was performed, but using only as evidence the values associated with two variables: difficulty walking a block and self-rating of health. These nodes were chosen since they were the most relevant nodes (most central nodes using degree, betweenness, and closeness) according to an analysis based on a Markov network [17]. A similar analysis was performed using the nine most connected nodes in the Bayesian network. Finally, the FI by itself was also used to classify dead people.

All analyses were performed in R-studio (R-version 3.5.1) through the bnlearn package [48] to implement the structural learning process, with the bootstrap method and hc algorithm, and for evidence propagation. On the other hand, the pROC [43] package was used to obtain ROC curves and analyze sensitivity and specificity after evidence propagation.

## Results

The network associated with the frailty components and death is shown in Figure 1. Nodes are colored according to the following classification: Yellow-AS (Affective Status: No_Happy, Lone, No_energy, Depressed, Tired, and Effort); Light Blue-Symptoms (Anorexia, Bed, and Lost_weight); Black-Dead; Gray-SRH (Self-rated health: Health and H_change); Green-ADL-IADL (activities of daily living and Instrumental activities of daily living: In_out_chair, Stairs, Shop, Dress, Walk, Eat, Groom, Meals, Meds, Finance, and Toilet); Orange-Physical Health (Grip, Exercise, Lift, and Walk_out); Pink-Comorbidities (CLD, Stroke, Cancer, CHF, Arthritis, Heart_Attack, Diabetes, and High_BP); Navy Blue-Cognition (Memory). Arcs are colored according to the strength of the relationship as a color gradient from yellow (less strength) to dark blue (more strength). We can see that all variables are connected, except Cancer, which is an isolated vertex, since there are no edges with enough strength considering the cut-off values used. Thus, Cancer is marginally independent to the other variables.

**Figure 1.**
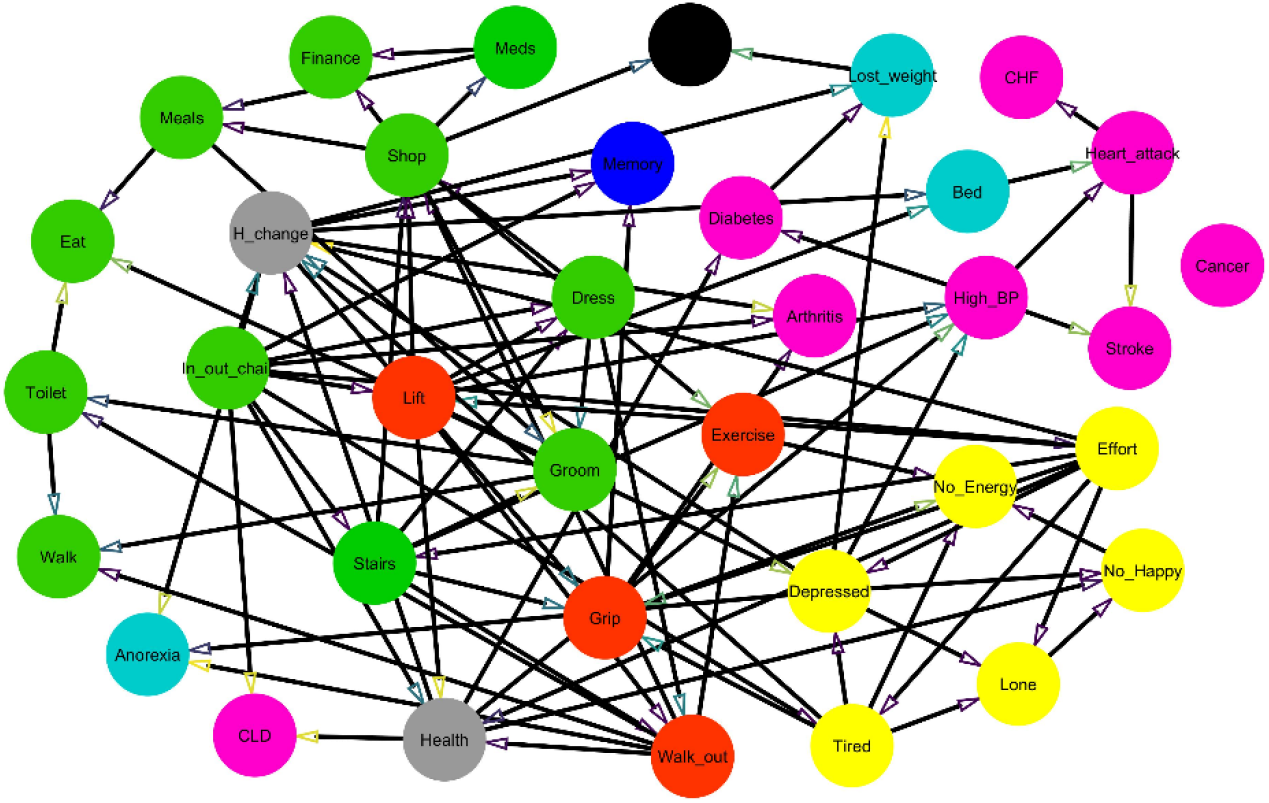
Averaged Bayesian network associated with frailty components and the dead node considering a Bootstrap sample of 100, using a hc algorithm with BIC score for each sample. Nodes are colored according to the following classification: Yellow-AS (Affective Status: No_Happy, Lone, No energy, Depressed, Tired, and Effort); Light Blue-Symptoms (Anorexia, Bed, and Lost_weight); Black-Dead; Gray-SRH (Self-rated health: Health and H_change); Green-ADL-IADL (activities of daily living and Instrumental activities of daily living: In_out_chair, Stairs, Shop, Dress, Walk, Eat, Groom, Meals, Meds, Finance, and Toilet); Orange-Physical Health (Grip, Exercise, Lift, and Walk_out); Pink-Comorbidities (CLD, Stroke, Cancer, CHF, Arthritis, Heart_Attack, Diabetes, and High_BP); Navy Blue-Cognition (Memory). A color scale distributes edge colors according to strength of the relationship between the variables: yellow-green-purple (from 0.5 to 1.0).

We observe that only two deficits are directly linked with the Dead node: Lost_weight and Shop, being Shop the node with the strongest relationship with Dead with a value of 0.85, whereas the value for Lost_weight is 0.69. There are four nodes pointing to Shop; the variable with the strongest relationship with Dead, which are Stairs, Dress, Lift, and Walk_out; thus, the probability associated with the Dead variable conditional to values provided to other variables not only depends on the Shop node. In fact, the ancestral set of the Dead node, nodes from which there are directed paths to Dead, is formed by 14 nodes (Figure 2).

**Figure 2.**
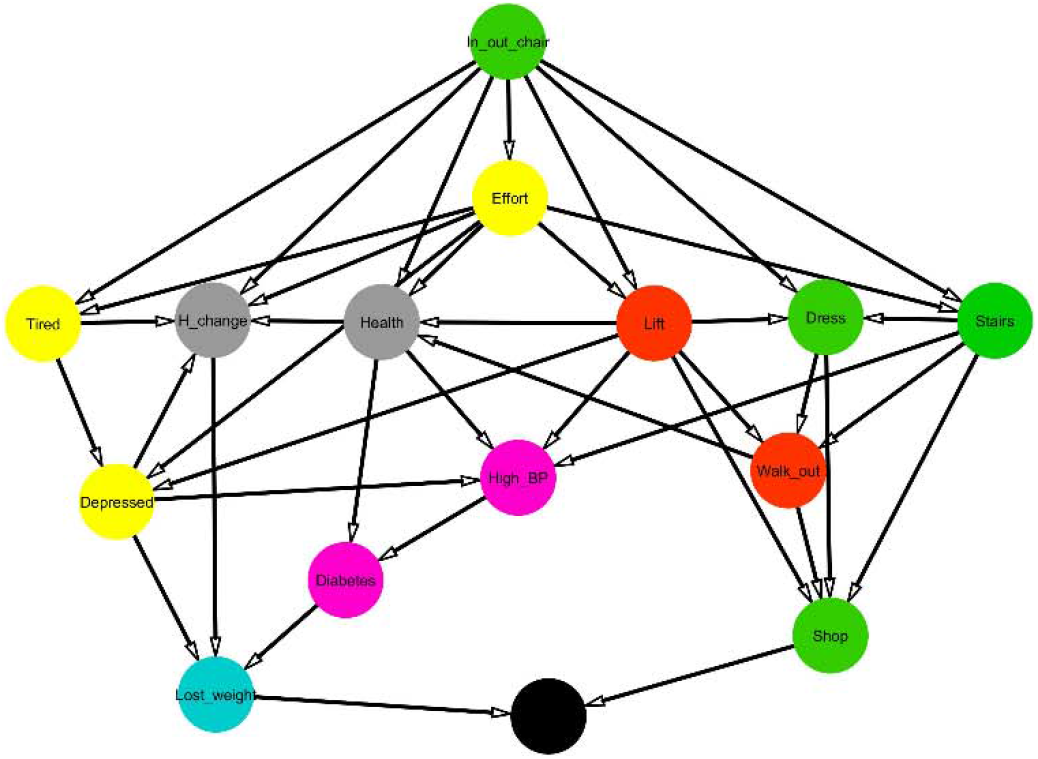
Directed graph induced by the ancestral set of the Dead node (nodes from which there are directed paths to Dead). Yellow-AS (Affective Status: Depressed, Tired, and Effort); Light Blue-Symptoms (Lost_weight); Black-Dead; Gray-SRH (Self-rated health: Health and H_change); Green-ADL-IADL (activities of daily living and Instrumental activities of daily living: In_out_chair, Stairs, Shop, and Dress); Orange-Physical Health (Lift and Walk_out); Pink Comorbidities (Diabetes and High_BP).

We observe that the variables with the greatest outdegree, with more arcs from the variable to any other variable, are In_out_chair with a value of ten, and Lift and Effort with a value of eight. On the other hand, the variables with the greatest indegree, with more arcs pointing to them, are Grip and H_change. It is noticeable that In_out_chair has an indegree of zero, though it has the greatest outdegree of all variables. When considering both arcs that begin or end into a variable, the variables with the greatest number of arcs are H_change with eleven arcs; In_out_chair, Lift, and Shop with ten arcs each one; and Health, Walk_out, Effort, Depressed, and Grip with nine arcs (Table 1).

**Table 1.**
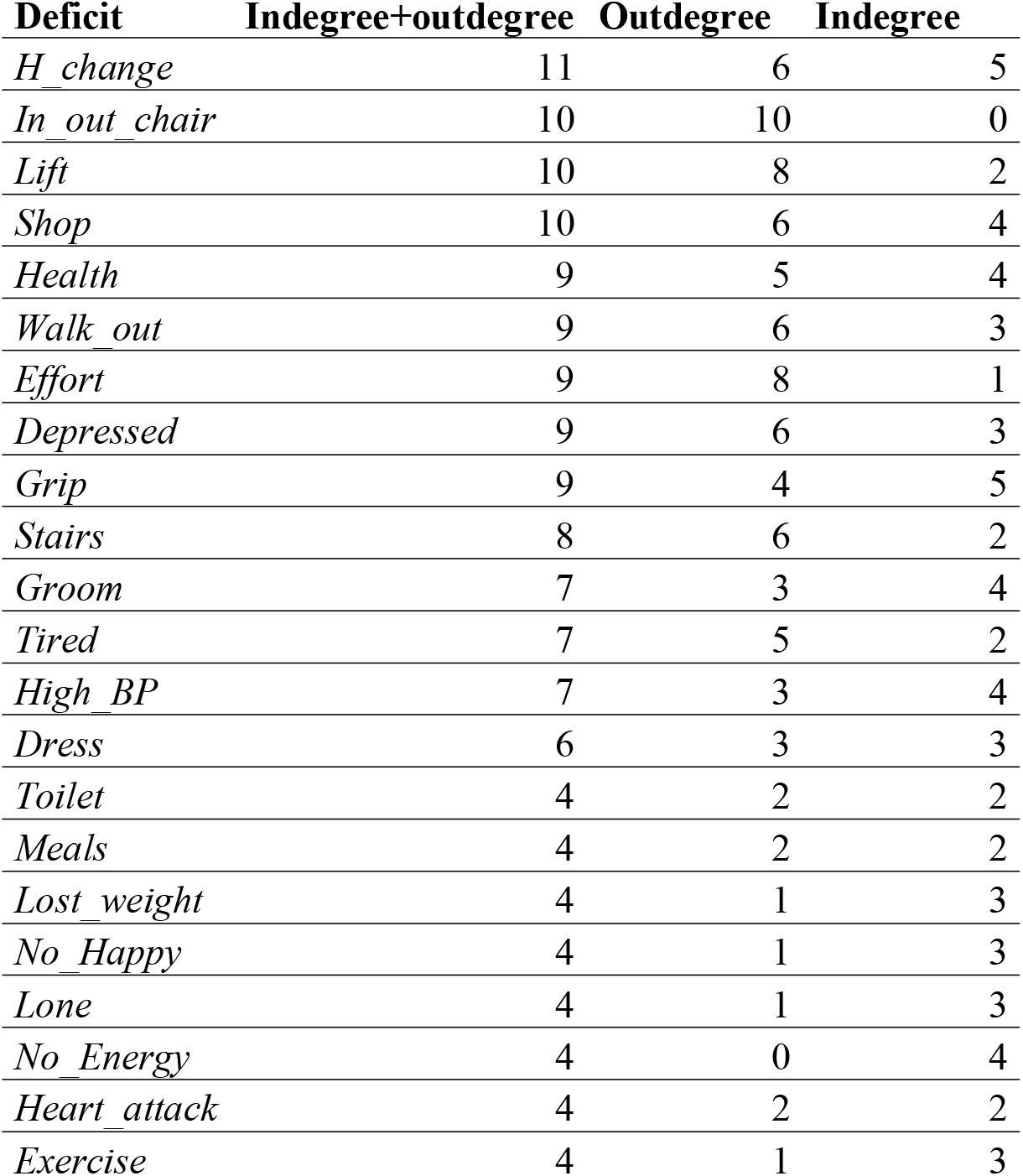
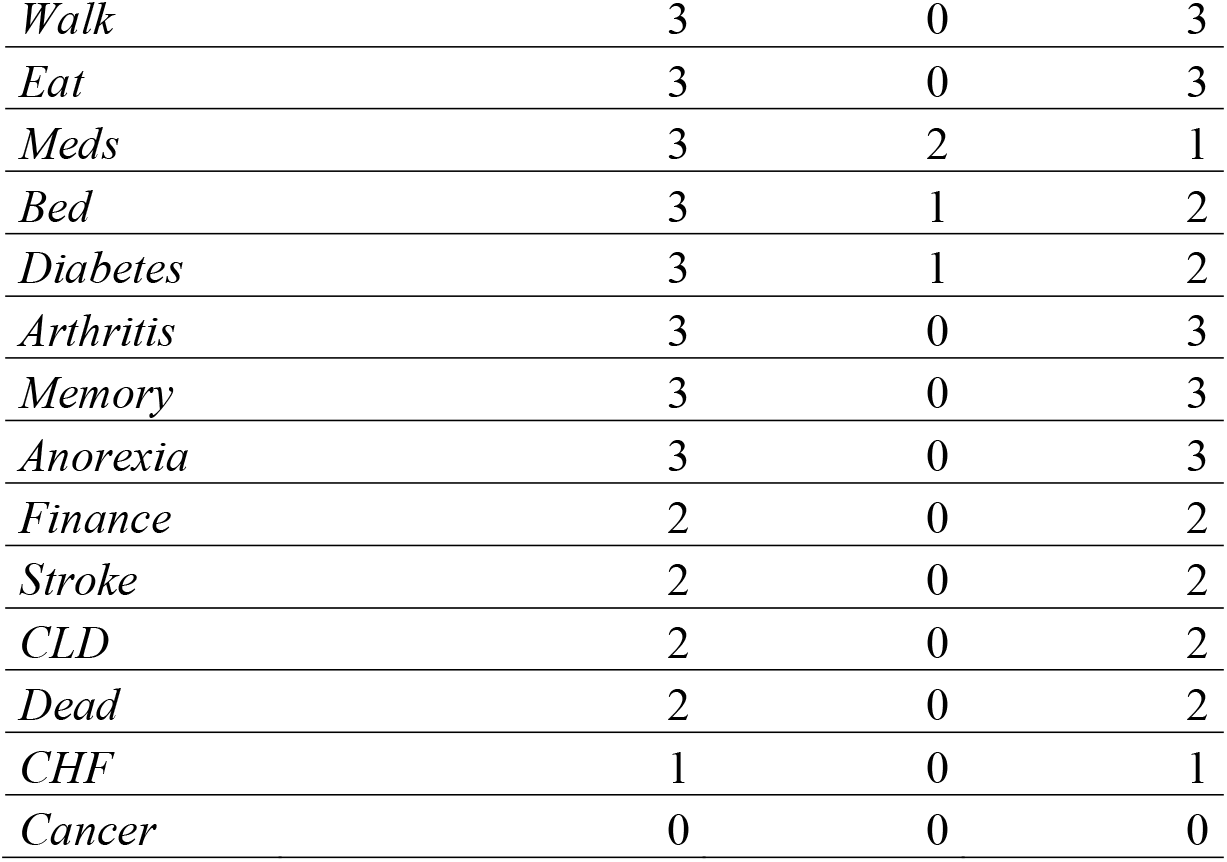
Indegree, outdegree, and sum of both measures for the averaged Bayesian network associated with frailty components and dead.

When considering the arcs with the highest strength, greater or equal to 0.8 for both the presence of an arc and its direction (Figure 3 and Supplementary material, Table 2), we observe direct relationships from the following nodes: From Depressed to Anorexia, Lone, and No_Happy; from Diabetes to Lost_weight; from Effort to Health, Depressed, and Lone; from Grip to Arthritis and Memory; from Groom to Toilet; from H_change to Bed and Memory; from Health to No_Happy, Diabetes and H_change; from Heart_attack to CHF; from High_BP to Diabetes and Heart_attack; from In_out_chair to Health, Memory, and Arthritis; from Lift to High_BP and Grip; from Meals to Eat; from Meds to Finance; from Shop to Dead, Meds, Finance, and Meals; from Stairs to Grip; from Tired to Depressed; from Lone to No energy; and from Walk_out to Toilet and Walk. Of course, there are indirect relationships, for instance from Effort to No_Happy trough the Depressed node.

**Figure 3.**
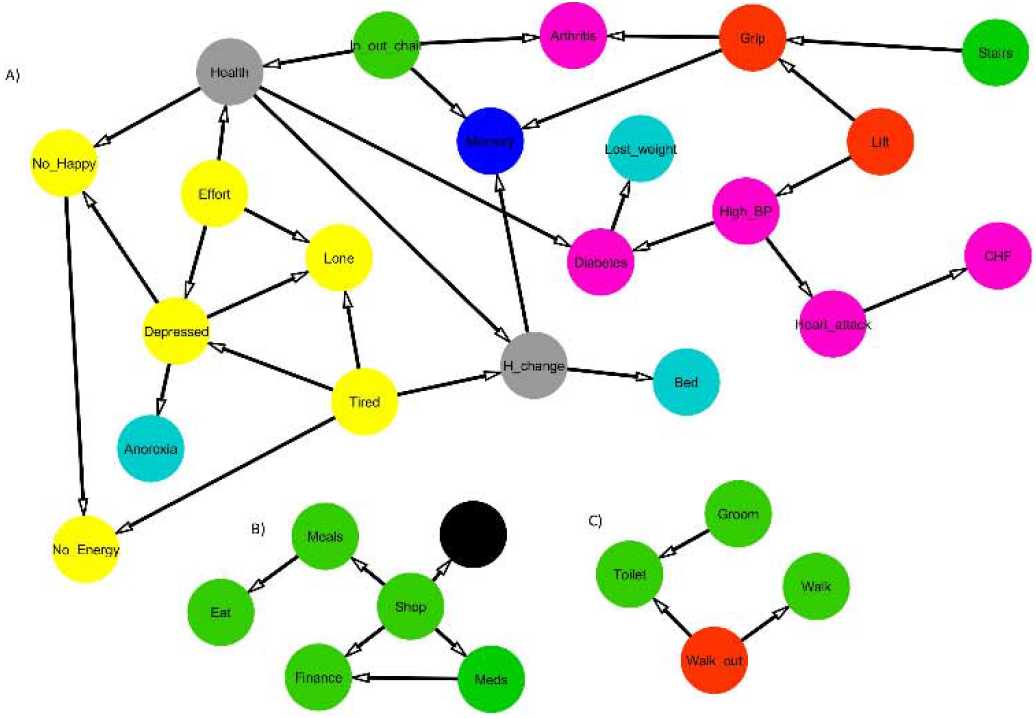
Network associated with the averaged Bayesian network for the frailty components and dead node induced by those arcs with a strength (both presence and direction) of 0.8 and more. Yellow-AS (Affective Status: No Happy, Lone, No energy, Depressed, Tired, and Effort); Light Blue-Symptoms (Anorexia, Bed, and Lost_weight); Black-Dead; Gray-SRH (Self-rated health: Health and H_change); Green-ADL-IADL (activities of daily living and Instrumental activities of daily living: In_out_chair, Stairs, Shop, Walk, Eat, Groom, Meals, Meds, Finance, and Toilet); Orange-Physical Health (Grip, Lift, and Walk_out); Pink Comorbidities (CHF, Arthritis, Heart_Attack, Diabetes, and High_BP); Navy Blue-Cognition (Memory). A, B, C correspond to the three connected components associated with the network.

After evidence associated with all nodes, except Dead, is propagated over the Dead node, we obtained the probability of death under the model for every individual. The corresponding ROC curve is shown in Figure 4 a) with an Area Under the Curve (AUC) of 0.6732. A new cut-off for assigning an individual to the dead class of 0.051 was obtained (individuals with probability greater or equal to that number are assigned to the dead category), obtaining through this cut-off value a sensitivity (death correctly assigned) of 56.1% and specificity of 74.6% and total percentage of correct assignation or accuracy of 73.58%.

**Figure 4.**
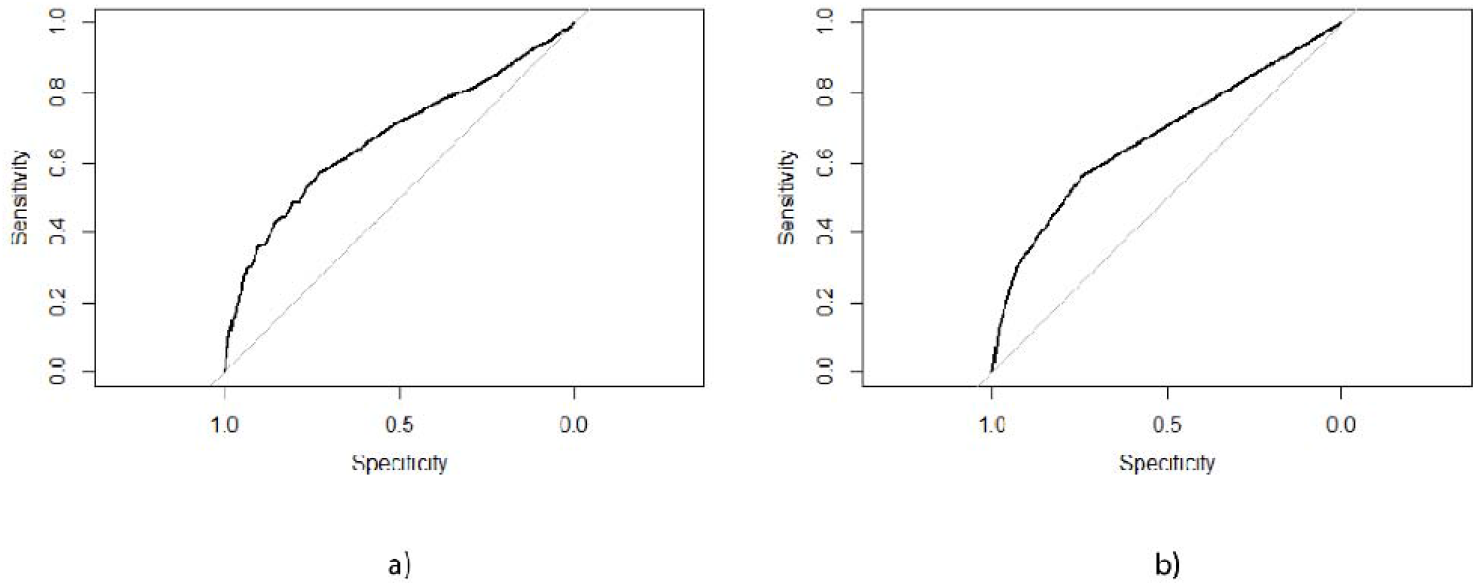
a) ROC curve model for evidence consisting of the values of all variables except dead; b) ROC curve model for evidence consisting of the values of variables Walk_out and Health.

When the values associated with the Walk_out and Health nodes are used as evidence and this evidence is once again propagated over the Dead node, we obtain the probability of death under the model and evidence provided by the two nodes. The associated ROC curve under the evidence is shown in Figure 4 b) with an AUC of 0.63. A new cut-off of 0.061 is obtained with an associated sensitivity of 37.98% and specificity of 84.76% and accuracy of 82.31%. When the values associated with the nine most connected nodes in the network in this paper are used (H_change, In_out_chair, Lift, Shop, Health, Walk_out, Effort, Depressed, and Grip), the AUC of the ROC curve (not shown) is the same, 0.63, with a cut-off value of 0.055, sensitivity and specificity of 38.15% and 87.27%, respectively, and accuracy of 84.7%, obtaining almost the same numbers as those obtained above using two nodes.

By only using the FI to separate between dead and alive people, being FI the average number of deficits by individual, and following a similar process of obtaining the associated ROC curve and optimum cut-off value, we obtained a AUC of 0.707, slightly better than the values obtained before, a cut-off value of 0.214 with an associated sensitivity of 60.97% and specificity of 68.90% and a accuracy of 68.49%

## Discussion

In the present study, we obtained a Bayesian network associated with frailty components and death, observing that not all nodes are similarly related between them, some have more arcs originated from them than others (outdegree), the same happened when considering arcs pointing to them (indegree). Interestingly, these unequal relationships were also found in our previous paper [17]. Additionally, through the network, we identified arcs, or in other words probabilistic causality, with more plausibility to be present, those arcs having the greatest strength. This indicates which relationships between the frailty components are more likely to occur and which nodes have a direct impact on death. In this work, we also provided values to all variables, except death, to calculate the probability of death for each individual and analyzed whether alive and death people can be correctly classified from using these variables. The same process was repeated using the values corresponding to other variables as evidence, and the results were compared.

As expected, variables of the same type are more likely to be linked together. For instance, there are arcs between comorbidities; from Heart_atack to CHF and Stroke and from High_BP to Diabetes, Hearth_Atack, and Stroke. Similarly, there are arcs between Affective Status (AS); for instance, from Depressed to Lone and No_Happy; from Effort to Depressed, Lone, and Tired; from Lone to No_Happy, and from the latter to No_Energy; and finally, from Tired to Depressed, Lone and No_Energy. There are also arcs between ADL and IADLs; for instance, from Meds to Meals and Finance. There are also arcs linking physical activities, one from Grip to Exercise, and others from Lift to Grip and to Walk_out, and another linking self-report of health, from Health to Health_change. On the other hand, there are arcs between variables of different types, for instance, there are arcs pointing to symptoms, for instance from Diabetes (comorbidity) and Depressed (AS) to Lost_weight or from Depressed (AS) to Anorexia. There are also arcs from Self-report of health variables to comorbidities and symptoms, for instance from Health to CLD, Diabetes, and High_BP (comorbidities) and from H_change to Anorexia, Bed, and Lost_weight (symptoms). Similarly, there are AS variables linked to physical health, for instance from Lift to Depressed. These allow us to perceive the complexity of the relationships within all components forming the frailty index.

As stated before, some nodes seem to be more relevant than others, and such importance should be considered in the index calculation, since they are all taken together as if they have the same impact. Apart from the highest number of connections (indegree and outdegree and their sum) and the analysis of arcs with the highest strength to be present and in its direction, the identification of nodes that are connected to death can be used to determine the most relevant items conforming the FI.

The node with the highest number of arcs (in or out) is report of current health compared to two years ago. Global health status, as measured by self-assessment, is a robust predictor of mortality and chronic morbidity, and is a very inclusive measure of health reflecting aspects relevant to survival which are not covered by other health indicators [23-24, 33, 59].

Functional impairment has long been an essential measure of disability in aging studies and in clinical practice to assess care needs. It is commonly measured using Activities of Daily Living (ADLs) and Instrumental Activities of Daily Living (IADLs). ADLs functions are essential for an individual’s self-care, whereas IADLs functions are more concerned with self-reliant functioning in each environment [18].

From the directed graph induced by the ancestral set of the Dead node (Figure 2), we identified that the node corresponding to difficulty getting in or out of a chair, which is an ADL, has the highest outdegree of all the variables and it is the initial node of all the directed trajectories to the Dead node. This feature agrees with the fact that the act of transferring is basic and critical in ADLs [51]. Many of the ADLs and IADLs such as eating, toileting, shopping, cooking, require an initial act of transfer to/from a sitting position before the performance of the activity. This importance is recognized in many scales including the Barthel Index [34] and its modifications [49] whereby a higher weightage is given to transfers compared to the other ADLs. In other instruments, such as the Functional Independence Measure [19], there are 13 motor items, of which there are three types of transfers: transfer to bed/chair, transfer to a shower/bathtub, and transfer to a toilet; and hence the ADL ‘transfer’ is triple the weightage of other ADLs.

On the other hand, several studies have shown that self-rating of health is a strong predictor of mortality, especially in the elderly [23, 26, 38], therefore, it is not surprising that this node belongs to the ancestral set of the Dead node. The importance of self-rated health for morbidity has also been studied as its relation to mental health, chronic cardiovascular diseases, and diabetes [4, 12, 21], which are major contributors to morbidity and mortality worldwide. Hence, as expected, depression, high blood pressure, and diabetes also belong to the ancestral set of the Dead node.

High blood pressure has long been recognized as an independent risk factor for fatal and nonfatal vascular events [32], leading to heart disease, stroke, and kidney failure, and if left uncontrolled, hypertension can lead to a heart attack, an enlargement of the heart, and eventually heart failure [6, 61]. In Mexico, cardiovascular disease is the most important cause of death; it is responsible for approximately 162 thousand deaths a year, nearly 23% of the total. Of these, complications of high blood pressure account for more than 80 thousand deaths every year [25]. Therefore, as seen in Figure 1, there were arcs from high blood pressure to heart attack and stroke. Similarly, heart attack was linked to chronic heart failure, which can be understood since all participants who reported CHF in this study, also suffered a heart attack [2].

Diabetes is also associated with an increased risk of complications as retinopathy, kidney disease, limb amputation, heart disease, and stroke [7, 27]. In Mexico, diabetes is responsible for 9% of deaths each year [25]. High blood pressure and diabetes are associated with each other, but whether a direct causal effect links the two is unclear. Some studies have shown that diabetes is causal for hypertension, but the impact of hypertension on the development of diabetes has provided conflicting findings [11, 54, 60]. Interestingly, according to our network, there is an arc from high blood pressure to diabetes with strength above 0.8. Hypertension and diabetes have etiological aspects in common [37], and often co-exist in the same individual [15], diabetes is more frequent in hypertensive than normotensive subjects [9, 20, 60]. For our study, 3252 individuals reported having only hypertension, 1475 reported having both conditions, and 919 participants reported having only diabetes. Therefore, there is a rationale to suspect that hypertension may cause incident diabetes, but further investigation is needed.

As shown in Figure 2, depression, worsening health status in the last two years, and diabetes are linked to unintentional weight loss (Lost_weight). Unintentional weight loss is common among elderly people and is associated with adverse outcomes and age-related diseases such as sarcopenia [1]. Among several physiologic, psychological, and social risk factors for involuntary weight loss, depression, and intake of medications for cardiac and endocrine illnesses have been associated to it [52].

Only Shop and Lost_weight point directly to the Dead node (Figures 1 and 2). Since there are several arcs pointing to these two variables, this does not mean that they only determine the probabilities associated with death when we provide values to other variables. However, probabilistically, it means that the Dead variable is conditionally independent to all other variables except Shop and Lost_weight (non-descendants) given Shop and Lost_weight, in other words, the other variables are linked to Dead through Shop and Lost_weight. Considering the relationship between weight loss and death, unintentional weight loss may reflect disease severity or undiagnosed illness [1]. Weight loss of 4%–5% or more of body weight within one year, or 10% or more over 5–10 years or longer, is associated with increased mortality or morbidity or both [1, 52]. In frail elderly populations, even small weight loss (e.g., 1 kg [41], or 3% of body weight [58]), may be significant. The relationship between the Shop and Dead nodes is discussed below.

As seen in Figure 3, when considering the arcs with the highest strength, greater or equal to 0.8 for both the presence of an arc and its direction, three clusters emerge. In cluster B, all the nodes linked with Dead (directly or indirectly) are IADLs (nodes in the arcs going from difficulty shopping to the nodes difficulty taking medications, managing finances, and preparing hot meals) and ADLs (nodes in the arc from difficulty preparing hot meals to difficulty eating).

In this sense, many ADLs and IADLs are conventionally scored by summing the responses to individual items on the scale to yield a total score. Despite the popularity of this method, some issues make it difficult to interpret [42], since the total-score method weights each item equally, which assumes that all items represent equal levels of severity. It has been suggested [35] that the eight items in the IADL scale could be placed on a hierarchy of ‘difficulty’ with ‘shopping’ the most discriminatory item differentiating well between patients of different levels of ability. ‘Shopping’, ‘food preparation’, and ‘medicine’ were found to be the most ‘difficult’ abilities to continue performing after a disease in patients with dementia and therefore those lost earliest. When ADLs are combined with IADLs, they better describe the spectrum of disability for a broader range of people [28] and it was also found that when combined [16, 55], ‘Shopping’ is placed second in the difficulty hierarchy (first place was taken by Do heavy cleaning or Cutting Toenails, which were not available in the data set we used to obtain the FI); thus, it makes sense its direct relationship with the Dead node.

Cluster C in Figure 3 relates difficulty walking a block (physical activity) with ADLs (nodes in the arcs going from the physical activity difficulty walking a block to difficulty using the toilet and walking around a room) and ADL’s (arc going from difficulty grooming to difficulty using the toilet). Clusters B and C contain 8 of the 11 ADLs and IADLs included in the FI.

On the other hand, Cluster A, which is the largest, contains two ADLs: difficulty getting in/out of a chair (which is the node with the highest outdegree, linked with 10 nodes) and difficulty up/down one flight of stairs. The node corresponding to difficulty dressing is the only one of the included ADLs and IADLs not appearing within the arcs with the highest strength (though it is weakly linked to Groom, Shop, Stairs, In_out_chair, Lift, and Walk_out). It is also noticeable that in this cluster the self-rated health nodes (Health and H_change) connect the affective status nodes with all the rest (i.e., in the underlying graph of the digraph, Health and H_change form a set of vertices separating affective status from the other nodes).

Surprisingly, Cancer was marginally independent to all other variables. When analyzing the network structure before considering any threshold; i.e. before obtaining the average network, there were arcs from and to Cancer. In particular, the arc with the highest strength, for both presence and direction, was from help dressing (Dress) to Cancer with values of 0.4 and 0.925, respectively. Thus, if we had used a threshold of 0.4 for the presence of an arc, it would have appeared. In fact, the direct association between Cancer and Dead was quite low, though significant (Pearson or Phi correlation of 0.060, p-value <0.001), thus indicating that perhaps at most an indirect relationship with the Dead node could exist, even though perhaps this relationship exists through variables not considered for our analyses.

When considering evidence provided by all nodes except Dead, people that died, was more correctly assigned using all nodes than when only two nodes, Walk_out and Health, or even the nine most connected nodes, were used. This means that the information provided by all deficits is important to identify the characteristics that people that died had.

However, since the sensitivity obtained through our model is 56%, it means that we are not completely characterizing what separates death from alive people, at least using the chosen deficits, which could mean that there are other important deficits that could be taken on account, though this could be for our data set only. It is noticeable that when the FI by itself is used as a classifier, the sensitivity increases almost in 5%, which is surprising considering that it is just an average, but FI has proved to be successful in this matter when compared with other models [29, 40, 53]. This does not mean that the FI is the best available classifier to distinguish between dead and alive people, but that with the defined deficits and in our population, the FI had slightly better results in terms of sensitivity. Even so, a classifier identifying barely 56% or 60% of dead individuals can seem not as good as desired, and in any case, a better classifier considering other aspects is needed. In this sense, if we wanted to create an improved classifier, we probably should include other type of variables, e..g socio-economic, molecular, or demographic, and in terms of Bayesian networks, maybe generate arcs guided more closely by experts instead of depending on algorithms, and work in the framework of supervised learning (creation of classifiers), i.e. using training and test sets, comparing with different classification models, using k-fold validation methods, etc. This analysis is out of the scope of this work and corresponds to further research. On the other hand, the relationships and relevance of certain nodes in terms of their connection and probabilistic causality, identified in this and our previous paper, could serve to improve the FI by modifying it as a weighted average, but this also requires further analyses.

Using samples concerning different populations, subpopulations (e.g. men and women), and time could also help to a better understanding of the real phenomenon, not only in terms of which causal relationships are or are not present between deficits in different populations, but also in terms of which variables composing the FI should be used when trying to separate dead and alive people. This is also future work.

One limitation of our work is that we are not considering possible censoring as in survival analysis. We are only considering survival at a fixed time point (2015), thus we are not considering that death occurs in different time points and that survival time could add relevant information. Additionally, all variables are self-reported, and thus some information could be inaccurate. Finally, we used a group of variables as components forming the FI, those recommended by the creators of the index; however, if we had used others, we do not know whether results could have been different.

## Conclusion

By using averaged Bayesian networks, we were able to identify causal relationships between deficits conforming the FI. We derived that not all deficits are similarly related, for instance report of current health compared to two years ago was the most connected node and difficulty getting in or out of a chair was the node with the greatest number of arcs parting from it and the node from which all directed trajectories to the *Dead* node start. When considering the strength of the relationships derived from a resampling process, arcs with a strength in their presence of 0.8 and over, we identified three clusters of nodes, two of them mostly involving ADL and IADL’s. Only two nodes point directly to the *Dead* node, difficulty shopping and weight loss, though there are several directed trajectories joining some nodes (14 nodes) to the Dead node. All these results seem plausible from a clinical perspective. As a classifier, dead people is identified in at most 61%, independently of the use or not of the Bayesian network with evidence propagation and the variables used in the model, suggesting that the FI is not as good classifier, at least in our data set, even though the FI has been found to be a relevant to explain mortality, being this feature a different task than to classify. The latter suggests other variables should be considered to obtain a better classifier, improving the correct classification of dead people. Finally, it might be useful to fit similar models in other populations and subpopulations to see whether the results are consistent.

## Data Availability

All data used are available online at http://mhasweb.org/Data.aspx
The derivation of each variable is explained in the Supplementary material

