## Supplementary tables for "Understanding frailty: probabilistic causality between components and their relationship with death through a Bayesian network and evidence propagation"

**Supplementary material. Appendix 1. Syntaxis for each variable**

| **Variable Description (name)** | **MHAS Variable** | **Coding of the variable** | **Syntaxis to generate variables** |
| --- | --- | --- | --- |
| Help dressing (Dress) | h13_12 Because of health problem, difficulty dressing self | 1 Yes  2 No  6 Can't do  7 Doesn't do  8 RF | Dress = h13_12_d_2  1 Yes if h13_12==1 \| h13_12==6 \| h13_12==7  0 No if h13_12==2 |
| Help getting in/out of a chair (In_out_chair) | h5_12 Because of health problem, difficulty getting up from chair | 1 Yes  2 No  6 Can't do  7 Doesn't do  8 RF | In_out_chair = h5_12_d_2  1 Yes if h5_12==1 \| h5_12==6 \| h5_12==7  0 No if h5_12==2 |
| Help walking around the room (Walk) | h15a_12 Because of health problem, difficulty walking | 1 Yes  2 No  6 Can't do  7 Doesn't do  8 RF  9 DK | Walk = h15d_12_d  1 Yes if (h15a_12==1 \| h15a_12==6 \| h15a_12==7) & h15d_12==1  0 No if h1_12==2 & h4_12==2 & h5_12==2 & h8_12==2 & h9_12==2 & h10_12==2 & h11_12==2 & h12_12==2 & h13_12==2 OR h15a_12==2 OR h15d_12==2 |
|  | h15d_12 Someone help you walk across room | 1 Yes  2 No  8 RF  9 DK |  |
| Help eating (Eat) | h17a_12 Because of health problem, difficulty eating or cutting | 1 Yes  2 No  6 Can't do  7 Doesn't do  8 RF  9 DK | Eat = h17d_12_d  1 Yes if (h17a_12==1 \| h17a_12==6 \| h17a_12==7) & h17d_12==1  0 No if h1_12==2 & h4_12==2 & h5_12==2 & h8_12==2 & h9_12==2 & h10_12==2 & h11_12==2 & h12_12==2 & h13_12==2 OR h17a_12==2 OR h17d_12==2 |
|  | h17d_12 Does someone help you eat your food | 1 Yes  2 No  8 RF  9 DK |  |
| Help grooming (Groom) | h16a_12 Because of health problem, difficulty bathing | 1 Yes  2 No  6 Can't do  7 Doesn't do  8 RF  9 DK | Groom = h16d_12_d  1 Yes if (h16a_12==1 \| h16a_12==6 \| h16a_12==7) & h16d_12==1  0 No if h1_12==2 & h4_12==2 & h5_12==2 & h8_12==2 & h9_12==2 & h10_12==2 & h11_12==2 & h12_12==2 & h13_12==2 OR h16a_12==2 OR h16d_12==2 |
|  | h16d_12 Someone  help you to bathe or shower | 1 Yes  2 No  8 RF  9 DK |  |
| Help using toilet (Toilet) | h19a_12 Because of health problem, difficulty going to the bathroom | 1 Yes  2 No  6 Can't do  7 Doesn't do  8 RF  9 DK | Toilet = h19d_12_d  1 Yes if (h19a_12==1 \| h19a_12==6 \| h19a_12==7) & h19d_12==1  0 No if h1_12==2 & h4_12==2 & h5_12==2 & h8_12==2 & h9_12==2 & h10_12==2 & h11_12==2 & h12_12==2 & h13_12==2 OR h19a_12==2 OR h19d_12==2 |
|  | h19d_12 Does someone help you use toilet, get on off | 1 Yes  2 No  8 RF  9 DK |  |
| Help up/down 1 flight of stairs (Stairs) | h6_12 Because of health problem, difficulty with flights of stairs | 1 Yes  2 No  6 Can't do  7 Doesn't do  8 RF  9 DK | Stairs = h7_12_d_2  h7_12_d=1 if h7_12==1  h7_12_d=0 if h6_12==2 \| h7_12==2  1 Yes if h7_12_d==1 \| h7_12==6  0 No if h7_12_d==0 |
|  | h7_12 Because of health problem, difficulty with 1 flight of stairs | 1 Yes  2 No  6 Can't do  7 Doesn't do  8 RF  9 DK |  |
| Help lifting 10 pounds (Lift) | h11_12 Because of health problem, difficulty carrying objects | 1 Yes  2 No  6 Can't do  7 Doesn't do  8 RF  9 DK | Lift = h11_12_d_2  1 Yes if h11_12==1 \| h11_12==6  0 No if h11_12==2 |
| Help shopping groceries (Shop) | h27a_12 Difficulty shopping | 1 Yes  2 No  6 Can't do  7 Doesn't do  8 RF  9 DK | Shop = h27c_12_d_2  h27c_12_d=1 if h27c_12==1  h27c_12_d=0 if h27a_12==2 \| h27c_12==2  1 Yes if h27c_12_d==1 \| h27a_12==6 \| h27a_12==7  0 No if h27c_12_d==0 |
|  | h27c_12 Does anyone help you shop for groceries | 1 Yes  2 No |  |
| Help with preparing hot meal (Meals) | h26a_12 Difficulty preparing hot food | 1 Yes  2 No  6 Can't do  7 Doesn't do  8 RF | Meals = h26c_12_d_2  h26c_12_d=1 if h26c_12==1  h26c_12_d=0 if h26a_12==2 \| h26c_12==2  1 Yes if h26c_12_d==1 \| h26a_12==6 \| h26a_12==7  0 No if h26c_12_d==0 |
|  | h26c_12 Does anyone help you prepare a hot meal | 1 Yes  2 No  8 RF  9 DK |  |
| Help taking medication (Meds) | h28a_12 Difficulty taking medications | 1 Yes  2 No  6 Can't do  7 Doesn't do  8 RF | Meds = h28c_12_d_2  h28c_12_d=1 if h28c_12==1  h28c_12_d=0 if h28a_12==2 \| h28c_12==2  1 Yes if h28c_12_d==1 \| h28a_12==6 \| h28a_12==7  0 No if h28c_12_d==0 |
|  | h28c_12 Does anyone help you take medications | 1 Yes  2 No |  |
| Help with finances (Finance) | h29a_12 Difficulty managing money | 1 Yes  2 No  6 Can't do  7 Doesn't do  8 RF  9 DK | Finance = h29c_12_d_2  h29c_12_d=1 if h29c_12==1  h29c_12_d=0 if h29a_12==2 \| h29c_12==2  1 Yes if h29c_12_d==1 \| h29a_12==6 \| h29a_12==7  0 No if h29c_12_d==0 |
|  | h29c_12 Does anyone one help you manage your money | 1 Yes  2 No  8 RF  9 DK |  |
| Compared to 2 years ago: Unintentional change in weight (Lost_weight) | c64_12 Compared to 2 years ago: Respondent's change in weight | 1 Has increased 5 kilos/more  2 Has decreased 5 kilos/more  3 Has remained more/less the same  8 RF  9 DK | Lost_weight = c64_12_d  1 Yes if c64_12==2 & c65_12==2  0 No if c64_12==1 \| c64_12==3 |
|  | c65_12 Compared to 2 years ago: Respondent's change in exercise or diet | 1 Yes  2 No  8 RF  9 DK |  |
| Self-rating of health (Health) | c1_12 Global self-reported quality of health | 1 Excellent  2 Very good  3 Good  4 Fair  5 Poor  8 RF  9 DK | Health = c1_12_d  1 Yes if c1_12==5  0 No if c1_12==1 \| c1_12==2 \| c1_12==3 \| c1_12==4 |
| Compared to 2 years ago: Report your current health (H_change) | c2a_12 Compared to 2 years ago: Report your current health | 1 Much better  2 Somewhat better  3 More or less the same  4 Somewhat worse  5 Much worse  9 DK | H_change = c2a_12_d  1 Yes if c2a_12==4 \| c2a_12==5  0 No if c2a_12==1 \| c2a_12==2 \| c2a_12==3 |
| Last 12 months: Number of days in bed... due to sickness/injury (Bed) | c73_12 Last 12 months: Respondent's number of days in bed... due to sickness/injury | | Bed = c73_12_d  1 Yes if c73_12>=1 & c73_12<=365  0 No if c73_12==0 |
| Within the past week: Felt tired (Tired) | c49_8_12 Within the past week: Respondent felt tired | 1 Yes  2 No  8 RF  9 DK | Tired = c49_8_12_d  1 Yes if c49_8_12==1  0 No if c49_8_12==2 |
| Difficulty walking a block (Walk_out) | h1_12 Because of health problem, difficulty walking blocks | 1 Yes  2 No  6 Can't do  7 Doesn't do  8 RF  9 DK | Walk_out = h3_12_d_2  h3_12_d=1 if h3_12==1  h3_12_d=0 if h1_12==2 \| h3_12==2  1 Yes if h3_12==6 \| h3_12==7 \| h3_12_d==1  0 No if h3_12_d==0 |
|  | h3_12 Because of health problem, difficulty walking a block | 1 Yes  2 No  6 Can't do  7 Doesn't do  8 RF  9 DK |  |
| Feel everything is an effort (Effort) | c49_2_12 Within the past week: Respondent experienced difficulty performing | 1 Yes  2 No  8 RF  9 DK | Effort = c49_2_12_d  1 Yes if c49_2_12==1  0 No if c49_2_12==2 |
| Feel depressed (Depressed) | c49_1_12 Within the past week: Respondent was depressed | 1 Yes  2 No  8 RF  9 DK | Depressed = c49_1_12_d  1 Yes if c49_1_12==1  0 No if c49_1_12==2 |
| Feel happy (No_happy) | c49_4_12 Within the past week: Respondent was happy | 1 Yes  2 No  8 RF  9 DK | No_happy = c49_4_12_d  1 Yes if c49_4_12==2  0 No if c49_4_12==1 |
| Feel lonely (Lone) | c49_5_12 Within the past week: Respondent was lonely | 1 Yes  2 No  8 RF  9 DK | Lone = c49_5_12_d  1 Yes if c49_5_12==1  0 No if c49_5_12==2 |
| Feel energetic (No_energy) | c49_9_12 Within the past week: Respondent was energetic | 1 Yes  2 No  8 RF  9 DK | No_energy = c49_9_12_d  1 Yes if c49_9_12==2  0 No if c49_9_12==1 |
| Hypertension/high blood pressure (High_BP) | c4_12 Has a physician diagnosed hypertension/high blood pressure | 1 Yes  2 No  8 RF  9 DK | High_BP = c4_12_d  1 Yes if c4_12==1  0 No if c4_12==2 |
| Heart attack (Heart_attack) | c22a_12 Has a physician ever told respondent heart attack | 1 Yes  2 No  8 RF  9 DK | Heart_attack = c22a_12_d  1 Yes if c22a_12 ==1  0 No if c22a_12 ==2 |
| Heart failure (CHF) | c25b_12 Has a physician ever told respondent heart failure | 1 Yes  2 No  9 DK | CHF = c25b_12_d  1 Yes if c25b_12 ==1  0 No if c22a_12==2 \| c25b_12==2 |
| Stroke (Stroke) | c26_12 Ever/last 2 years: Has a physician told respondent stroke | 1 Yes  2 No  8 RF  9 DK | Stroke = c26_12_d  1 Yes if c26_12 ==1  0 No if c26_12 ==2 |
| Cancer (in the last 10 years) (Cancer) | c12_12 Has a physician diagnosed respondent cancer  (last 10 years) | 1 Yes  2 No  8 RF  9 DK | Cancer = c12_12_d  1 Yes if c12_12 ==1 last 10 years  0 No if c12_12 ==2 last 10 years |
| Diabetes (Diabetes) | c6_12 Has a physician diagnosed respondent diabetes | 1 Yes  2 No  8 RF  9 DK | Diabetes = c6_12_d  1 Yes if c6_12 ==1  0 No if c6_12 ==2 |
| Arthritis/rheumatism (Arthritis) | c32_12 Has a physician diagnosed respondent with arthritis/rheumatism | 1 Yes  2 No  8 RF  9 DK | Arthritis = c32_12_d  1 Yes if c32_12 ==1  0 No if c32_12 ==2 |
| Respiratory illness (CLD) | c19_12 Has a physician diagnosed respondent respiratory illness | 1 Yes  2 No  8 RF  9 DK | CLD = c19_12_d  1 Yes if c19_12 ==1  0 No if c19_12 ==2 |
| Compared to 2 years ago: respondent reports his/her memory quality (Memory) | e1b_12 Compared to 2 years ago: respondent reports his/her memory quality | 1 Better  2 About the same  3 Worse  8 RF  9 DK | Memory = e1b_12_d  1 Yes if e1b_12==3  0 No if e1b_12<=2 |
| Respondent's dominant hand strength (Grip) | c69a_12 Respondent's dominant hand strength | 1 Very strong  2 Somewhat strong  3 Somewhat weak  4 Very weak  8 RF  9 DK | Grip = c69a_12_d  1 Yes if c69a_12==3 \| c69a_12==4  0 No if c69a_12<=2 |
| Anorexia: Last 2 years: Loss of appetite (Anorexia) | c70_12 Last 2 years: Respondent's loss of appetite | 1 Often  2 Sometimes  3 Rarely  8 RF  9 DK | Anorexia = c70_12_d  1 Yes if c70_12==1  0 No if c70_12==2 \| c70_12==3 |
| Exercise: Last 2 years: Exercise or hard physical work >= 3 times a week (Exercise) | c50b_12 Last 2 years: Respondent exercise or hard physical work >= 3 times a week | 1 Yes  2 No  8 RF  9 DK | Exercise = c50b_12_d  1 Yes if c50b_12==2  0 No if c50b_12==1 |
| Died between 2012 and 2015 (Dead) | fallecido_15 Died between 2012 and 2015 | 1 Yes  0 No | Dead  1 Yes if fallecido_15==1  0 No if fallecido_15==0 |
| *Filter participants with 50 or more years old  Variable for age 2012 is age_12, then age_12_cat=1 if age_12>=50, age_12_cat=0 if age_12<50  Filter: age_12_cat==1 | | | |
| **Eliminate second or third wives or husbands  Variable for subject identification is np, np = 10 original subject, np = 20 original spouse  Filter: np==10 \| np==20 | | | |
| ***The following variables have the same coding:  h1_12 Because of health problem, difficulty walking blocks  h4_12 Because of health problem, difficulty staying seated  h5_12 Because of health problem, difficulty getting up from chair  h8_12 Because of health problem, difficulty sitting up  h9_12 Because of health problem, difficulty lifting arms  h10_12 Because of health problem, difficulty pushing or pulling  h11_12 Because of health problem, difficulty carrying objects  h12_12 Because of health problem, difficulty picking up a coin  h13_12 Because of health problem, difficulty dressing self | | | 1 Yes  2 No  6 Can't do  7 Doesn't do  8 RF  9 DK |

**Supplementary material. Appendix 2.**

| Arc from | Arc to | Strength (Presence) | Strength (Direction) | Arcs with greatest strength (value =1 if presence and direction >=0.8) |
| --- | --- | --- | --- | --- |
| Depressed | Anorexia | 0.91 | 1.00 | 1 |
| Depressed | Lone | 1.00 | 0.94 | 1 |
| Depressed | No_Happy | 1.00 | 0.99 | 1 |
| Diabetes | Lost_weight | 1.00 | 0.99 | 1 |
| Effort | Depressed | 1.00 | 0.85 | 1 |
| Effort | Health | 0.93 | 0.85 | 1 |
| Effort | Lone | 1.00 | 0.94 | 1 |
| Grip | Arthritis | 0.99 | 0.99 | 1 |
| Grip | Memory | 0.99 | 0.97 | 1 |
| Groom | Toilet | 0.87 | 0.91 | 1 |
| H_change | Bed | 0.86 | 1.00 | 1 |
| H_change | Memory | 1.00 | 1.00 | 1 |
| Health | Diabetes | 1.00 | 0.96 | 1 |
| Health | H_change | 1.00 | 0.99 | 1 |
| Health | No_Happy | 0.97 | 0.91 | 1 |
| Heart_attack | CHF | 1.00 | 1.00 | 1 |
| High_BP | Diabetes | 1.00 | 0.99 | 1 |
| High_BP | Heart_attack | 1.00 | 1.00 | 1 |
| In_out_chair | Arthritis | 1.00 | 1.00 | 1 |
| In_out_chair | Health | 0.81 | 0.90 | 1 |
| In_out_chair | Memory | 0.99 | 1.00 | 1 |
| Lift | Grip | 1.00 | 0.98 | 1 |
| Lift | High_BP | 0.84 | 0.99 | 1 |
| Meals | Eat | 0.95 | 0.98 | 1 |
| Meds | Finance | 1.00 | 0.94 | 1 |
| No_Happy | No_Energy | 1.00 | 1.00 | 1 |
| Shop | Dead | 0.85 | 1.00 | 1 |
| Shop | Finance | 1.00 | 0.97 | 1 |
| Shop | Meals | 1.00 | 0.84 | 1 |
| Shop | Meds | 0.89 | 0.89 | 1 |
| Stairs | Grip | 0.81 | 0.98 | 1 |
| Tired | Depressed | 1.00 | 0.84 | 1 |
| Tired | H_change | 0.80 | 0.95 | 1 |
| Tired | Lone | 1.00 | 0.94 | 1 |
| Tired | No_Energy | 1.00 | 1.00 | 1 |
| Walk_out | Toilet | 0.94 | 0.98 | 1 |
| Walk_out | Walk | 0.99 | 0.95 | 1 |
| Bed | Heart_attack | 0.68 | 1.00 | 0 |
| Depressed | H_change | 0.81 | 0.62 | 0 |
| Depressed | High_BP | 0.76 | 0.99 | 0 |
| Depressed | Lost_weight | 0.52 | 1.00 | 0 |
| Dress | Groom | 0.79 | 0.89 | 0 |
| Dress | Shop | 0.99 | 0.55 | 0 |
| Dress | Walk_out | 0.79 | 0.65 | 0 |
| Effort | Grip | 0.69 | 0.97 | 0 |
| Effort | H_change | 0.51 | 0.97 | 0 |
| Effort | Lift | 0.75 | 0.47 | 0 |
| Effort | Stairs | 0.99 | 0.50 | 0 |
| Effort | Tired | 1.00 | 0.51 | 0 |
| Exercise | No_Energy | 0.99 | 0.77 | 0 |
| Grip | Exercise | 0.62 | 1.00 | 0 |
| Grip | No_Energy | 0.62 | 0.90 | 0 |
| Groom | Eat | 0.64 | 0.98 | 0 |
| Groom | Walk | 0.84 | 0.54 | 0 |
| H_change | Anorexia | 0.57 | 1.00 | 0 |
| H_change | Arthritis | 0.57 | 1.00 | 0 |
| H_change | Grip | 0.80 | 0.78 | 0 |
| H_change | Lost_weight | 0.77 | 1.00 | 0 |
| Health | CLD | 0.54 | 0.98 | 0 |
| Health | High_BP | 0.67 | 1.00 | 0 |
| Heart_attack | Stroke | 0.57 | 0.64 | 0 |
| High_BP | Stroke | 0.63 | 1.00 | 0 |
| In_out_chair | CLD | 0.54 | 0.95 | 0 |
| In_out_chair | Dress | 1.00 | 0.67 | 0 |
| In_out_chair | Effort | 0.96 | 0.52 | 0 |
| In_out_chair | H_change | 0.79 | 0.99 | 0 |
| In_out_chair | Lift | 1.00 | 0.60 | 0 |
| In_out_chair | Stairs | 1.00 | 0.53 | 0 |
| In_out_chair | Tired | 1.00 | 0.54 | 0 |
| Lift | Bed | 0.75 | 0.99 | 0 |
| Lift | Depressed | 0.61 | 0.62 | 0 |
| Lift | Dress | 0.99 | 0.64 | 0 |
| Lift | Health | 0.54 | 0.84 | 0 |
| Lift | Shop | 1.00 | 0.70 | 0 |
| Lift | Walk_out | 1.00 | 0.69 | 0 |
| Lone | No_Happy | 1.00 | 0.51 | 0 |
| Lost_weight | Dead | 0.69 | 1.00 | 0 |
| Meals | Groom | 0.85 | 0.58 | 0 |
| Meds | Meals | 0.95 | 0.19 | 0 |
| Shop | Exercise | 0.66 | 0.94 | 0 |
| Shop | Groom | 0.54 | 0.86 | 0 |
| Stairs | Dress | 1.00 | 0.67 | 0 |
| Stairs | Groom | 0.56 | 0.92 | 0 |
| Stairs | High_BP | 0.78 | 0.99 | 0 |
| Stairs | Shop | 0.96 | 0.69 | 0 |
| Stairs | Walk_out | 1.00 | 0.70 | 0 |
| Tired | Grip | 0.75 | 0.96 | 0 |
| Toilet | Eat | 0.59 | 0.98 | 0 |
| Toilet | Walk | 0.81 | 0.17 | 0 |
| Walk_out | Anorexia | 0.52 | 1.00 | 0 |
| Walk_out | Exercise | 0.70 | 0.93 | 0 |
| Walk_out | Health | 0.98 | 0.75 | 0 |
| Walk_out | Shop | 1.00 | 0.65 | 0 |

**Arcs and strength, for both the presence and direction, corresponding to the averaged Bayesian network concerning frailty components and death.** The arcs with the greatest strength, greater or equal to 0.8, for both the presence and direction of arcs, correspond to the first 37 (last column with value of one). The colors in the first two columns represent the classification of nodes: Yellow-AS (Affective Status); Light Blue-Symptoms; Red-Dead; Gray-SRH (Self-rated health); Green-ADL-IADL (activities of daily living and Instrumental activities of daily living); Orange-Physical Health; Pink-Comorbidities CLD; Navy Blue-Cognition; and the colors in the two last two columns represent presence of a node strength of 0.8 and more in yellow and in blue strength of a direction only above of 0.8 and more)
